# Amyloid-Independent Elevation of Plasma phosphorylated tau-217 Tracks Neurodegenerative Risk in Tuberous Sclerosis Complex: Implications for mTOR-Targeted Intervention

**DOI:** 10.64898/2026.09.09.26362677

**Authors:** Jocelyn Baumel, Michael Lutz, Cynthia Ding, Jamie Capal, David Ritter, Darcy Krueger, Alice Salter, Klaus Werner, Andy J. Liu

**Affiliations:** Department of Pediatrics, Division of Pediatric Neurology and Developmental Medicine, Duke University School of Medicine, 3000 Erwin Rd., Ste 100, Durham, NC 27705, USA; Department of Neurology, Division of Translational Brain Sciences, Duke University School of Medicine, 311 Research Drive, Durham, NC 27710, USA; Department of Neurology, Division of Behavioral Neurology, Duke University School of Medicine, 932 Morreene Rd., Durham, NC 27705, USA; Department of Neurology, Cincinnati Children’s Hospital Medical Center, 3333 Burnet Ave., Cincinnati, OH 45229, USA; Department of Pediatrics, University of Cincinnati College of Medicine, 3230 Eden Ave., Cincinnati, OH 45267, USA

**Keywords:** Tuberous Sclerosis Complex (TSC), Tuberous Sclerosis Complex Associated Neuropsychiatric Disorders (TAND), Alzheimer’s disease (AD), phosphorylated tau-217 (ptau-217)

## Abstract

**Background:** Tuberous sclerosis complex (TSC) is a rare autosomal dominant neurodevelopmental disorder caused by loss-of-function mutations in *TSC1* or *TSC2*, resulting in chronic hyperactivation of the mechanistic target of rapamycin (mTOR) pathway. TSC patients experience a wide spectrum of cognitive, behavioral, and psychiatric features collectively termed Tuberous Sclerosis Complex Associated Neuropsychiatric Disorders (TAND). Moreover, these patients develop a clinical trajectory in adulthood with clinical and neuropathological features overlapping those of Alzheimer’s disease (AD). Prior neuropathological studies have shown accumulation of AD-type mixed 3R/4R tau isoforms in adults with TSC that is amyloid-independent.

**Methods:** To further explore the biological convergence of TSC and AD, this study compared plasma levels of phosphorylated tau-217 (ptau-217), a sensitive and specific marker of early AD pathology, in 63 TSC, 25 AD, and 116 cognitively unimpaired (CU) patients.

**Results:** Findings revealed elevated levels of plasma ptau-217 in individuals with TSC at younger ages with a parallel age-related trajectory between TSC, AD, and CU cohorts. Multivariable linear regression showed that the log_2_ ptau-217 concentrations were significantly lower in the CU cohort relative to TSC (β = −0.21, SE = 0.05, p < 0.0001) but were not significantly different between the TSC and AD cohorts (β = 0.17, SE = 0.09, p < 0.071). The comparison of slopes analysis (ANCOVA) supported that the slopes for the regression of log_2_ ptau-217 versus age are not significantly different for the TSC and CU cohorts (TSC slope = 0.030, SE = 0.008; CU slope = 0.019, SE = 0.006; p = 0.26), nor for the TSC and AD cohorts (TSC slope = 0.030, SE = 0.01; AD slope = 0.06, SE = 0.03; p = 0.099).

**Conclusions:** These results suggest a novel link between TSC and AD that substantiates shared downstream neurodegenerative pathways. Furthermore, these results support the potential utility of ptau-217 as a minimally invasive biomarker for TAND severity, neurodegenerative risk, and AD-like prognosis in TSC and as a tool to guide new mTOR-targeted therapeutic strategies.

## Background

Tuberous sclerosis complex (TSC) is a rare autosomal dominant neurodevelopmental disorder caused by loss-of-function mutations in the *TSC1* or *TSC2* genes, which encode the proteins hamartin and tuberin (1). Together, these proteins form a complex that negatively regulates the mechanistic target of rapamycin (mTOR) signaling pathway (2). Disruption of this regulatory function leads to chronic mTOR hyperactivation, resulting in abnormal cellular growth, impaired homeostasis, and the development of benign tumors across multiple organ systems (1,3). Sarnat and Flores-Sarnat characterized TSC as an infantile tauopathy, with mTOR pathway activation affecting tau metabolism in immature neurons altering developmental processes (4). These alterations during development contribute to the formation of epileptogenic foci (4). Neurologic manifestations are central to TSC and extend beyond epilepsy.

Individuals with TSC commonly experience a wide spectrum of cognitive, behavioral and psychiatric symptoms collectively known as Tuberous Sclerosis Complex Associated Neuropsychiatric Disorders (TAND) (5).

TAND includes intellectual disability, autism spectrum disorder, attention-deficit/hyperactivity disorder, behavioral dysregulation, and mood disorders (5). Differences in neurodevelopment can be detected as early as 6 months of age with delays in fine motor and visual reception domains, followed by qualitative impairment in social communication and language delays as early as 9 to 12 months (6,7). In addition to these early-onset neurodevelopmental features, many individuals with TSC develop progressive impairments in memory, executive function, language, and behavior that interfere with educational attainment, occupational functioning, and quality of life. Notably, beginning in mid-to-late adulthood, a subset of individuals with TSC demonstrate cognitive decline that clinically overlaps with Alzheimer’s disease (AD) dementia (8–10).

Regarding the pathogenesis, chronic mTOR hyperactivation exerts broad downstream effects relevant to neurodegeneration, including impaired autophagy, synaptic dysfunction, oxidative stress, neuroinflammation, and abnormal protein aggregation (11). Converging evidence suggests that tau pathology may represent a shared biological substrate linking TAND and AD. Neuroimaging studies using tau-specific positron emission tomography radiotracers have demonstrated accumulation of flortaucipir (AV1451) in patients with TSC, a tracer with high affinity for phosphorylated tau 181 (ptau-181) linked to AD. This radioligand is well established to exhibit off-target binding (9, 12, 13). Subsequently, complementary neuropathological studies in adults with TSC showed sparse neurofibrillary tangles that stain with both AV1451 and GT-38. GT-38 is an antibody that detects mixed 3R/4R isoforms of tau seen in AD, in the absence of insoluble amyloid-β deposition (8). Cerebrospinal fluid analyses in TSC participants further demonstrated elevated ptau-181 and neurofilament light chain, consistent with ongoing neurodegeneration due to AD pathology (9).

Collectively, these findings support the classification of TSC as a distinct tauopathy, characterized by widespread neurofibrillary tangle deposition across cortical, limbic, and subcortical regions. Although these tangles share key biochemical features with AD, including mixed 3R/4R tau isoforms, they display distinct post-translational modifications, regional distributions, and an amyloid-independent pathogenesis (8,13). Experimental data further indicate that neurons with heightened mTOR activity— particularly hippocampal CA1 neurons, a region highly vulnerable in AD—are selectively predisposed to tau accumulation and degeneration, implicating mTOR dysregulation as a central upstream driver (14,15).

Within this framework, phosphorylated tau at threonine 217 (ptau-217) emerges as a promising biomarker. Ptau-217 is a sensitive and specific marker of early AD pathology, localizes to neurofibrillary tangles and neuropil threads, and correlates strongly with brain tau burden (16–20). Building on the mechanistic convergence between TSC and AD, we report elevated plasma ptau-217 levels in individuals with TSC compared with healthy controls. These findings highlight the potential utility of ptau-217 as a minimally invasive biomarker for TAND severity, neurodegenerative risk, and AD-like prognosis in TSC, as well as a tool to guide mTOR-targeted therapeutic strategies.

## Methods

### Plasma sample collection and handling

140 blood samples from 63 patients with TSC confirmed by genetic testing were obtained through the TSC Alliance Biosample Repository. Associated, deidentified data variables from the TSC Natural History Database including demographics (biological sex, race, ethnicity), cardiovascular disease, comorbidities (chronic infections, diabetes, autoimmune disease, etc.), clinical or molecular diagnosis, genetics, neurological disease (epilepsy, SEGA, SENs, tubers), neurological treatments including antiepileptic drugs, neuropsychiatric factors (intellectual ability, psychiatric disorders, learning disorders, behaviors), use of psychoactive drugs, other TSC conditions not captured elsewhere, and renal manifestations (angiomyolipoma, cysts, polycystic kidneys) were requested. Of note, chronic kidney disease status was requested but not obtained due to unavailability.

Blood samples from 116 cognitively unimpaired (CU) patients with CSF negative for AD biomarkers and 25 patients diagnosed with AD as defined by core 1 biomarker criteria from the National Institute on Aging and the Alzheimer’s Association work groups outlined by Jack et al. (21) were obtained through the Duke-University of North Carolina Alzheimer’s Disease Research Center (ADRC). All samples were shipped on dry ice and stored without preclinical complication.

### Plasma ptau-217 SIMOA measurements

Ptau-217 SIMOA measurements for the TSC were performed in a Quanterix HD-X instrumentation platform in the Molecular Genomics Core of the Duke Molecular Physiology Institute. Plasma ptau-217 measurements used high precision Quanterix SIMOA ptau-217 Advantage PLUS Reagent kit (cat # 104570; lot # 504576; Quanterix Corp., Billerica, MA, USA). Calibrators were run in triplicates, controls run in duplicates, and plasma ptau-217 SIMOA were measured in duplicates. All plasma samples were diluted on bench manually using vendor’s diluent and protocol.

In the AD and CU cohorts, plasma ptau-217 samples were run at the National Centralized Repository for Alzheimer’s and Related Disease (NCRAD) at Indiana University as part of the Duke-UNC Alzheimer’s Disease Research Center (ADRC) standard operating procedure. NCRAD also used the Quanterix HD-X platform to complete these results using the Alzpath ptau-217 kit. The vendor’s protocol was used to run these samples.

### Statistical analysis

Of the 140 blood samples from the TSC cohort, only the earliest collected sample for each patient was included for analysis. Descriptive statistics were calculated for participant characteristics, including clinical, demographic, genetic (TSC mutations), and ptau-217 biomarker data to provide a clear overview of the cohorts and highlight differences that might influence the results. For continuous variables (age), we conducted two sample t-tests to assess mean differences between cohorts (TSC, AD, CU). Categorical variables with two categories were tested for cohort differences using a two-tailed Fisher’s exact test. Ptau-217 measurements were log_2_ transformed to normalize the right-skewed distribution of the data and to stabilize variance.

The effect of TSC mutations on ptau-217 levels was examined using a multivariable linear model with log_2_ ptau-217 concentrations as the dependent variable, cohort (TSC, AD, CU) as the independent variable, and age and sex as covariates. The effect of the covariates was assessed by evaluation of effect size and statistical significance. Model fit was assessed by examination of residuals and leverage plots. The relationship between age and log_2_ ptau-217 concentrations stratified by cohort were analyzed using linear regression and ANCOVA comparison of slopes.

A secondary analysis based on a sample based on propensity score matching was performed to mitigate the differences between the number of individuals in the TSC, AD, and CU cohorts and adjust for potential confounders including age and sex. Adequacy of the propensity score matching was assessed by examination of standardized mean difference. Matching ratio was 1:1 using the greedy nearest neighbor algorithm without replacement and a caliper setting of 0.25. A standardized mean difference < 0.20 was considered acceptable covariate balance for the propensity score matching. The balance of covariates post-matching was examined. Multivariable linear regression to examine the effect of TSC mutations on ptau-217 levels was performed using the same approach as for analysis of the entire sample.

A two-sided p-value < 0.05 was considered statistically significant. All statistical procedures were performed using JMP version 19.0.1 (SAS Institute, Cary, NC).

## Results

Demographic, clinical, and ptau-217 biomarker data for the three cohorts are shown in Tables 1 and 2. In total, 204 patients were included in the study. 116 CU patients had CSF negative for AD biomarkers, 25 were diagnosed with AD, and 63 patients were diagnosed with TSC. The AD cohort was the eldest of the three, with age ranging from 51 to 77 and a mean age of 64.8 years. The TSC cohort’s age ranged from 41 to 79 years, with a mean age of 53.1 years. The CU cohort age ranged from 40 to 78 years, with a mean age of 57.4 years. The sex distribution across groups was similar with a slight female predominance in each cohort (56% female in the AD, 61.2% in the CU, and 61.9% in the TSC cohort). The CU cohort had a higher mean age (57.4 versus 53.1 years) and greater racial diversity with higher proportions of Black patients (12% versus 1.6%) compared to the TSC cohort.

**Table 1:** Demographics of the Study Cohorts by Diagnosis. Table 1 shows the demographic characteristics of patients in the CU, AD, and TSC cohorts. Age is presented as mean and standard deviation. Race and sex are presented as frequency and percentage.

|  | CU | AD | TSC | p (CU vs. TSC) | p (AD vs. TSC) |
| --- | --- | --- | --- | --- | --- |
| N | 116 | 25 | 63 |  |  |
| Age (Mean, SD) | 57.4 (9.7) | 64.8 (8.5) | 53.1 (9.9) | 0.006 | <0.0001 |
| Sex (N, percent female) | 71, 61.2% | 14, 56% | 39, 61.9% | 0.93 | 0.64 |
| Race (N, percent) |  |  |  | 0.08 | 0.66 |
| Black | 14, 12% | 0, 0% | 1, 1.6% |  |  |
| White | 97, 84% | 24, 96% | 59, 93.6% |  |  |
| Hispanic | 0, 0% | 0, 0% | 1, 1.6% |  |  |
| Asian | 1, 1% | 0, 0% | 1, 1.6% |  |  |
| Other | 4, 3% | 1, 4% | 1, 1.6% |  |  |

**Table 2:**
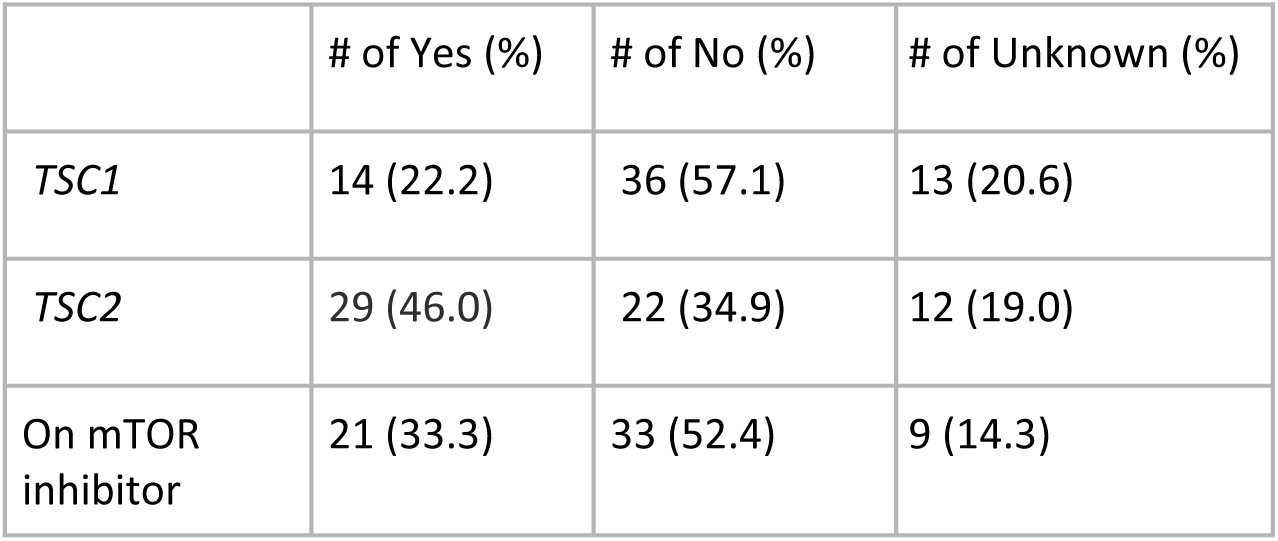
Clinical Profiles of TSC Cohort. Table 2 summarizes the genetic mutations of individuals in the TSC cohort and whether they were using an mTOR inhibitor.

|  | # of Yes (%) | # of No (%) | # of Unknown (%) |
| --- | --- | --- | --- |
| TSC1 | 14 (22.2) | 36 (57.1) | 13 (20.6) |
| TSC2 | 29 (46.0) | 22 (34.9) | 12 (19.0) |
| On mTOR inhibitor | 21 (33.3) | 33 (52.4) | 9 (14.3) |

Multivariable linear regression showed that the log_2_ ptau-217 concentrations were significantly different between the TSC and CU cohorts (β = −0.21, SE = 0.05, p < 0.0001, reference is TSC). Age was significantly associated with log_2_ ptau-217 concentrations (β = 0.02, SE = 0.005, p < 0.0001). Sex was not significantly associated with log_2_ ptau-217 concentrations (β = −0.03, SE = 0.05, p = 0.5, reference is male). Residual plots did not show systematic lack of fit by predicted Y values; leverage plots did not show systematic lack of fit by values of the covariates.

Multivariable linear regression showed that the log_2_ ptau-217 concentrations were not significantly different between the TSC and AD cohorts (β = 0.17, SE = 0.09, p = 0.071, reference is TSC). Age was significantly associated with log_2_ ptau-217 concentrations (β = 0.04, SE = 0.008, p < 0.0001); sex was not significantly associated with log_2_ ptau-217 concentrations (β = −0.07, SE = 0.07, p = 0.32, reference is male). Residual plots did not show systematic lack of fit by predicted Y values; leverage plots did not show systematic lack of fit by values of the covariates.

The association between log_2_ ptau-217 and age for the TSC, AD, and CU cohorts is shown in Figure 1. The comparison of slopes analysis (ANCOVA) supported that the slopes for the regression of log_2_ ptau-217 versus age are not significantly different for the TSC and CU cohorts (CU slope = 0.019, SE = 0.006; TSC slope = 0.030, SE = 0.008; p = 0.26), nor for the TSC and AD cohorts (AD slope = 0.06, SE = 0.03; TSC slope = 0.030, SE = 0.01; p = 0.099).

**Figure 1:**
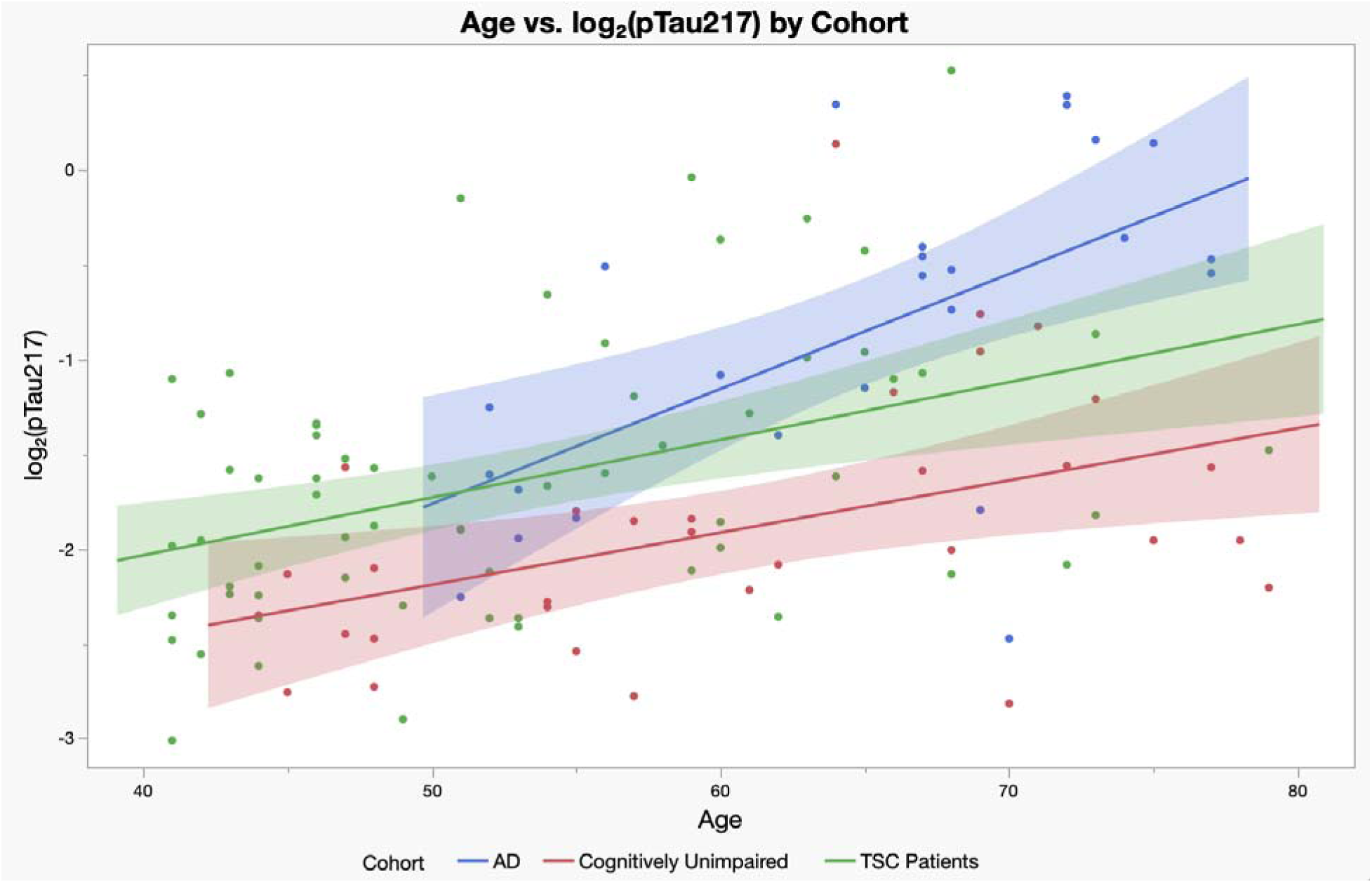
The Association Between Age and Log_2_ Ptau-217 by Cohort. Figure 1 shows the relationship between age and plasma log_2_-transformed ptau-217 concentrations in individuals from the TSC, AD, and CU cohorts. Solid lines represent fitted linear regressions while shaded areas indicate the 95% confidence interval.

Comparisons of cohort-specific slopes for the association between log_2_ ptau-217 and age for TSC versus CU and AD cohorts stratified by sex showed that the slopes were not significantly different for females (TSC versus CU: CU slope = 0.015, SE = 0.008; TSC slope = 0.027, SE = 0.009; p = 0.32; TSC versus AD: AD slope = 0.05, SE = 0.02; TSC slope = 0.03, SE = 0.01; p = 0.33) or for males (TSC versus CU: CU slope = 0.024, SE = 0.01; TSC slope = 0.035, SE = 0.02; p = 0.56; TSC versus AD: AD slope = 0.07, SE = 0.03; TSC slope = 0.04, SE = 0.02; p = 0.24). This comparison is shown in Figure 2.

**Figure 2:**
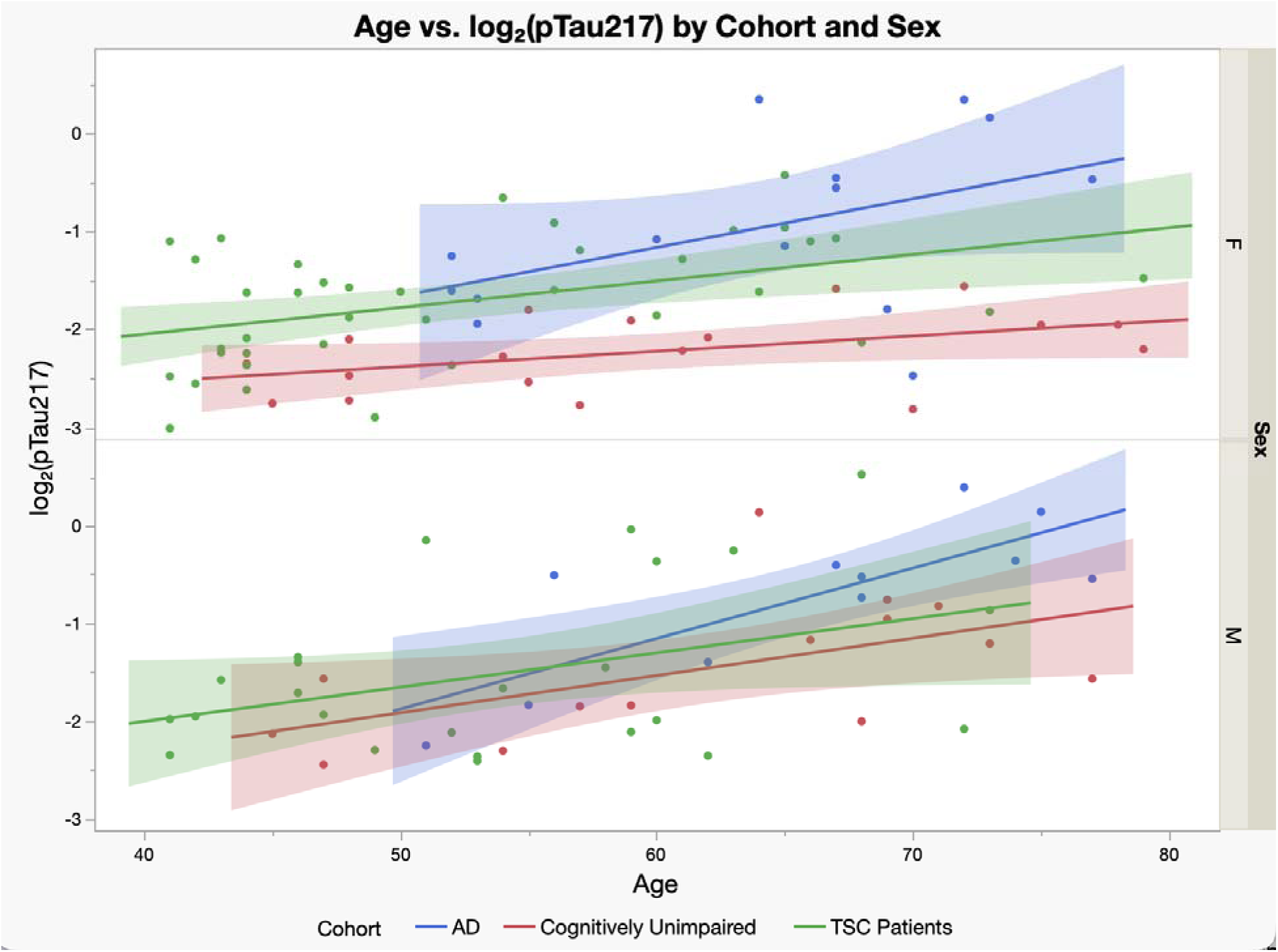
The Association Between Age and Log_2_ Ptau-217 by Cohort and Sex. Figure 2 shows the relationship between age and plasma log_2_-transformed ptau-217 concentrations, stratified by sex, in individuals from the TSC, AD, and CU cohorts. Solid lines represent fitted linear regressions while shaded areas indicate the 95% confidence interval.

For the propensity score matched analysis, the association between log_2_ ptau-217 and cohort (TSC versus CU) remained significant (β = −0.23, SE = 0.05, p < 0.0001). The propensity score matching based on age and sex shows a reduction in standardized mean difference from 0.27 to 0.15, a 42% reduction, supporting adequate balance for these variables. Examination of the covariates post-matching confirmed strong balance with mean age for the TSC group at 55.2 years (SD = 9.9) versus 53.6 years (SD = 9.8) for the CU group; sex was balanced at 60% female for both cohorts.

For the propensity score matched analysis, the association between log_2_ ptau-217 and cohort (TSC versus AD) remained not significant (β = 0.18, SE = 0.12, p = 0.14). The propensity score matching based on age and sex shows a reduction in standardized mean difference from 1.2 to 0.09, a 93% reduction, supporting adequate balance for these variables. Examination of the covariates post-matching confirmed strong balance with mean age for the TSC group at 62.8 years (SD = 8.1) versus 63.3 years (SD = 8.5) for the AD group; sex was balanced at 57% female for both cohorts.

## Discussion

To our knowledge, this is the first study to investigate plasma ptau-217 as a biomarker in individuals with TSC. Previous studies in TSC have focused primarily on alternative biomarkers including ptau-181 and GFAP, whereas plasma ptau-217 is well-established as a sensitive and specific marker of AD–related tau pathology (9,16–20,22). Moreover, this study is novel in comparing plasma ptau-217 levels between individuals with TSC and patients with AD.

Although TSC-associated tauopathy has classically been described in the absence of amyloid-β deposition (8,13), we observed elevated plasma ptau-217 levels in TSC, with an age-related trajectory also seen in AD (23,24). Notably, individuals with TSC demonstrated higher ptau-217 levels at younger ages compared to healthy controls, suggesting early emergence of AD-like tau changes. Furthermore, while sex-specific differences in plasma ptau-217 levels did not reach a level of significance, we observed a trend toward greater separation in confidence intervals between the CU versus the TSC and AD cohorts. This mirrors sex-specific patterns seen in p-tau biomarker studies in AD across multiple large cohorts which have demonstrated elevated plasma ptau-217 levels compared to men in the presence of high Aβ burden (25). These observations support a previously unrecognized link between TSC and AD and raise the possibility that cognitive decline in TSC may be more closely aligned with AD-related mechanisms than previously appreciated. Collectively, these findings suggest that TSC may represent a genetically mediated tauopathy with downstream features overlapping those of an amyloid-independent AD pathology.

In CU, amyloid-negative AD populations, correlations between plasma ptau-217 and age are typically modest or absent (correlation coefficients ranging from 0.48 to −0.02), consistent with limited age-driven change (16,23,24). In contrast, amyloid positivity in AD patients is associated with substantially accelerated increases in ptau-217, with prospective longitudinal studies demonstrating a six-fold higher annual increase in plasma ptau-217 in amyloid-positive versus amyloid-negative individuals (16,23,26). These changes are detectable in preclinical stages and predictive of progression to mild cognitive impairment (MCI) and AD dementia in a dose-dependent relationship (16,23,27). Plasma ptau-217 has repeatedly demonstrated strong prognostic performance and generally outperforms other plasma biomarkers in predicting conversion from MCI to dementia (20,24). The age-associated increase observed in our TSC cohort suggests that ptau-217 may have comparable utility for identifying individuals with TSC at heightened risk for cognitive decline. Future directions include collecting results of neuropsychologic testing to determine whether the relationship between ptau-217 levels and cognitive decline seen in AD holds true in the TSC population and whether a similar dose dependent relationship occurs with TAND severity.

Our prior neuropathological study on TSC brain tissue showed no amyloid plaque accumulation (8). The increase in plasma ptau-217 and absence of amyloid plaque in TSC brain tissue participants suggests that ptau-217 accumulation may occur independently of amyloid deposition. This also suggests that hyperactivation of the mTOR pathway is responsible for the accumulation of ptau-217.

Strengths of this study include the relatively large TSC sample and cohort matching. A power analysis from our previous work based on cerebrospinal fluid ptau-181 levels in TSC indicated that 40 participants would be sufficient to detect group differences (9); our study included 63 individuals with TSC. The three cohorts were well matched for age, sex, and race, reducing potential demographic confounding. The propensity score matched analysis further supports this. Moreover, the residual plots did not show a systemic lack of fit suggesting that our current statistical model is appropriate.

Several limitations warrant consideration. First, comorbid conditions—particularly chronic kidney disease (CKD)—may influence plasma tau concentrations. CKD, which is common in TSC, has been associated with increased plasma p-tau levels independent of age or AD pathology, likely reflecting reduced peripheral clearance rather than increased central production, as cerebrospinal fluid tau does not show parallel elevations (26,28). This may be contributing to the higher baseline level of ptau-217 seen in TSC patients when compared to CU patients in this study. Reported increases in p-tau with declining estimated glomerular filtration rate (eGFR) are modest relative to amyloid-associated changes, and plasma phosphorylated-to-total tau217 ratios substantially mitigate renal confounding (29). CKD prevalence in TSC ranges from approximately 12–13% in pediatric populations to 20–40% in adults, underscoring the importance of accounting for renal function in future biomarker studies (30–32). More specific ptau-217 named brain derived ptau-217 is now available which would further mitigate CKD as a confounding factor (33). Comparable rates of impaired kidney function have been reported in aging and AD research cohorts, and lower eGFR has not consistently correlated with cognitive impairment in these populations (34).

Secondly, although amyloid plaques were not seen in TSC brain tissue (8), soluble forms of amyloid could still be present. Cerebrospinal fluid analysis of Aβ42levels in TSC patients were found to be comparable to those of healthy controls making soluble forms of amyloid less likely (9).

Additional biological factors, including underweight status and anemia, have been reported to influence plasma ptau-217 independent of amyloid status (26). Elevated cerebrospinal fluid p-tau levels are also observed in frontotemporal dementia (FTD) associated with MAPT variants, and comparable CSF p-tau levels have been reported in TSC and FTD cohorts, suggesting that increased p-tau is not exclusively specific to AD pathology (9).

## Conclusion

Building on emerging evidence of mechanistic convergence between TSC and AD, this study demonstrates elevated plasma ptau-217 levels in individuals with TSC. Prior neuropathologic studies describe TSC as a distinct tauopathy characterized by predominant 3R/4R tau isoforms and absence of amyloid-β deposition further supporting the link between TSC and AD-related tau. These results support the hypothesis of partially overlapping neurodegenerative pathways and suggest that plasma ptau-217 may serve as a minimally invasive biomarker for TAND, neurodegenerative risk stratification, and AD-like prognostication in TSC. Additionally, pathogenic studies on the relationship between the mTOR pathway and plasma ptau-217 have the potential to facilitate the development of new biomarkers to diagnose TAND in TSC patients along with staging AD patients earlier in the disease course. Identifying mTOR pathway biomarkers also has the potential to discover new therapeutics such as FDA approved mTOR inhibitors for both TSC and AD patients.

## Abbreviations

Aβ42: amyloid β-42
AD A: lzheimer’s disease
ADRC: Duke/University of North Carolina Alzheimer’s Disease Research Center
CKD: chronic kidney disease
MCI: mild cognitive impairment
mTOR: mechanistic target of rapamycin
Ptau-181: phosphorylated tau-181
Ptau-217: phosphorylated tau-217
TAND: Tuberous Sclerosis Complex Associated Neuropsychiatric Disorders
TSC: Tuberous Sclerosis Complex

## Declarations

### Ethics approval and consent to participate

This study is in accordance with the Declaration of Helsinki and was approved by the Duke University Hospital System Institutional Review Board. Informed consent was waived due to the retrospective nature of the study.

### Consent for publication

Not applicable

### Availability of data and materials

The datasets used and analyzed during the current study are available from the corresponding author on reasonable request.

### Authors’ contributions

Study conception and design: AL. Data analysis: ML, CD. Drafting of the article: JB, CD, ML. Review and editing: AL, JC, DR, DK, AS, KW. All authors contributed to the article and approved the submitted version.

## Acknowledgements

The authors would like to thank all of the patients who have participated in donating plasma samples to the TSC Alliance and the Duke-UNC ADRC along with the medical teams providing their care.

